# Subtypes of early childhood caries predict future caries experience

**DOI:** 10.1101/2022.01.10.22268959

**Authors:** Alexander Gormley, Simon Haworth, Miguel Simancas-Pallares, Pernilla Lif Holgerson, Anders Esberg, Poojan Shrestha, Kimon Divaris, Ingegerd Johansson

## Abstract

**Objectives:** To test whether postulated subtypes of early childhood caries (ECC) are predictive of subsequent caries experience in a population-based cohort of Swedish children.

**Methods:** The study included children aged between 3 and 5 years at study entry with dental records available for at least 5 years of follow-up. Dental record data were retrieved from the Swedish Quality Registry for Caries and Periodontal disease (SKaPa) for the initial and follow-up visits. Participants who had ECC at study entry were assigned to one of five ECC subtypes (termed classes 1 to 5) using latent class modelling of tooth surface-level caries experience. Subsequent experience of caries was assessed using the decayed, missing, and filled surfaces indices (dmfs/DMFS) at follow-up visits, and compared between ECC subtypes using logistic and negative binomial regression modelling.

**Results:** The study included 128,355 children who had 3 or more dental visits spanning at least 5 years post baseline. Of these children, 31,919 had caries at the initial visit. Baseline ECC subtype was associated with differences in subsequent disease experience. As an example, 83% of children who had a severe form of ECC at age 5 went on to have caries in the permanent dentition by the end of the study, compared to 51% of children who were caries-free at age 5 (adjusted odds ratio of 4.9 for new disease at their third follow-up).

**Conclusions:** ECC subtypes assigned at a baseline visit are associated with differences in subsequent caries experience in both primary and permanent teeth. This suggests that the development and future validation of an ECC classification can be used in addition to current prediction tools to help identify children at high risk of developing new caries lesions throughout childhood and adolescence.

## 1 INTRODUCTION

Early Childhood Caries (ECC) is defined as the presence of one or more decayed (non-cavitated or cavitated), missing (due to caries) or filled tooth surfaces in any tooth in the primary dentition in a child 6 years and younger.^1^ It is an important disease of childhood as it affects the oral and general wellbeing of the child, in addition to the economic, educational, and generalised impairment to quality of life and costs this disease has on society.^2^ Sequalae of dental caries can include impaired eating, and educational disadvantage as well as the need for invasive treatment, for example under sedation or general anaesthesia.^3,4^ The global burden of ECC remains high despite a reduction in caries prevalence in recent decades.^5^ Thus, there remains a high burden of both restored and unrestored caries lesions in children, which is a major problem in the global context of non-communicable diseases of health.^6^

In the clinical setting, it would be useful to understand which children are at highest risk of caries. Past caries experience is a recognised predictor for new caries experience^7^, and caries experience in primary teeth can predict caries experience in the permanent dentition.^8^ This may be because children who develop caries at a young age have a high burden of genetic, environmental, or behavioural risk factors, and these same risk factors are important for both primary and permanent teeth. If so, clinically manifest disease is acting as a proxy measure for underlying risk factors. Or children are simply ‘disease active’ and this person-level disease activity continuously and progressively affects more tooth surfaces, as they emerge in the oral cavity. Recent data suggest the established definition of ECC does not reflect the heterogeneous clinical presentations of the disease. Newer subtypes are now postulated in ECC,^9^ and in adult caries,^10^ which could reflect variation in the underlying aetiology. If true, aetiological differences between these subtypes might result in different patterns of subsequent disease presentation, and different initial presentations of caries might proxy different underlying risk factors. ECC subtype classification might therefore be a useful tool in identifying children who are especially susceptible to new or progressive disease and help improve assessment, prediction and diagnosis of caries within paediatric, but also adult, populations. ^11^

If different presentations of disease represent different underlying processes, then characterizing the pattern of presentation should add diagnostic value^12^ in a clinical setting. With better understanding of the underlying molecular, genetic, microbiological and environmental risk factors underlying different diseases it may be possible to propose effective targeted interventions. This concept of personalised or precision dentistry has been discussed in the literature, with a goal towards delivering interventions personalised to the patient for the managed prevention and treatment of oral diseases,^12,13^ but to date there are few practical applications.

This study aimed to investigate whether ECC subtypes can predict variation in subsequent caries experience. If so, this could highlight a role for assigning caries subtypes in precision prevention and management.

## 2 METHODS

### 2.1 Study group

The study used a longitudinal design nested in a national Swedish register of dental records. From a basic cohort of 492,350 children with data from dental examinations from the Swedish Quality Registry for Caries and Periodontal Disease (SKaPa, www.skapareg.se), children between 3 and 5 years old at their first visit were retained (n=208,112 children) (Figure 1). SKaPa is a national register that started to compile data from dental examinations in 2008. Data are automatically extracted from electronic patient dental examination records from all public dental clinics in Sweden in addition to a growing number of private clinics^14^. In Sweden, free dental care is offered, and provided, to all children at the public care clinics and therefore the data from SKaPa are considered representative of the entire childhood population of Sweden. From this initial baseline cohort, participants were excluded if they did not have information on at least three follow-up visits spanning over 5 years. The final study group was 128,355 children (Figure 1).

**FIGURE 1.**
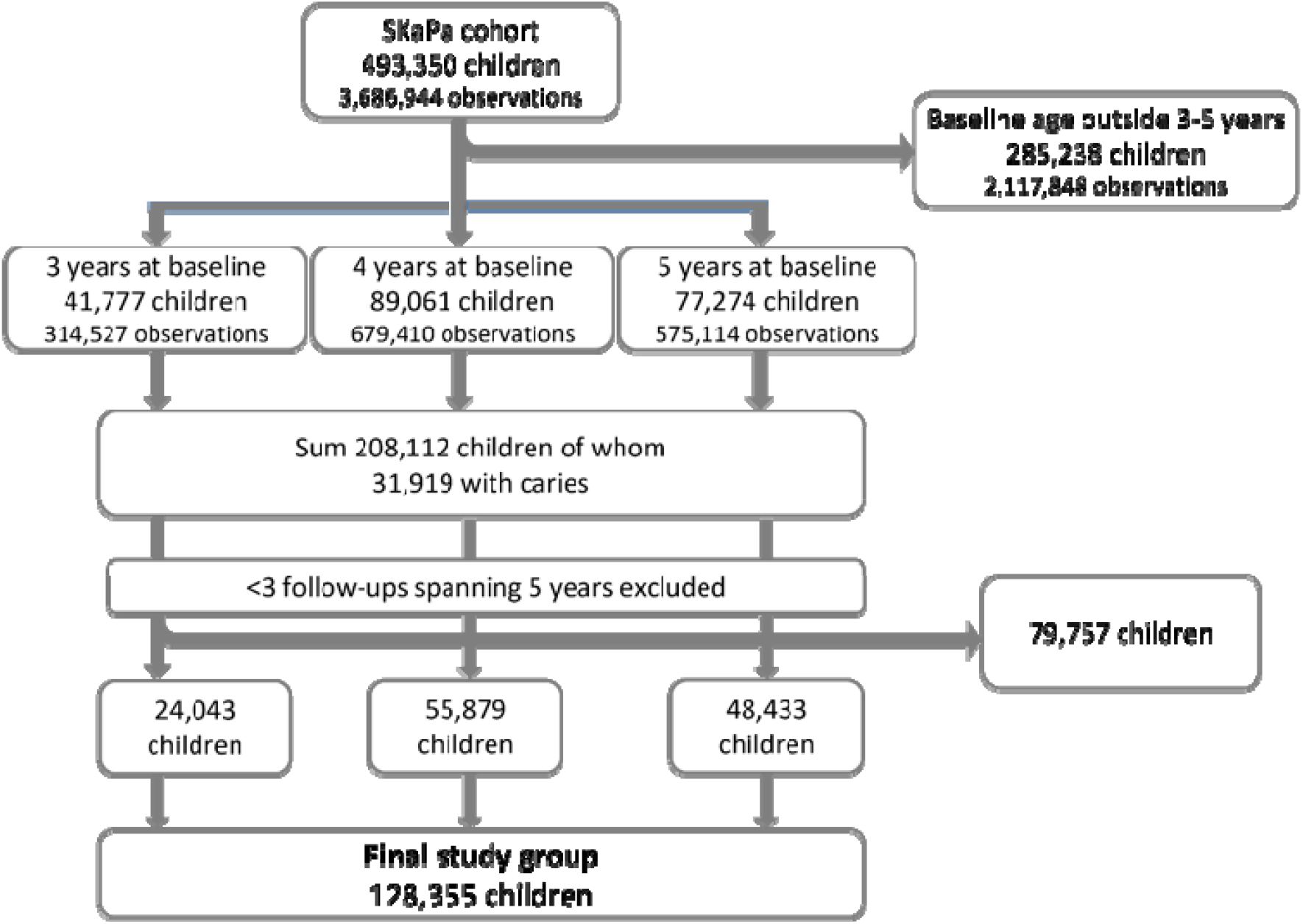
Flow chart of final analysis sample

### 2.2 Caries coding and analyses

Each tooth surface is charted separately and given a status code representing the condition of that tooth surface, as determined by the examining dentist. Status codes for each tooth surface (buccal, lingual, mesial, distal and occlusal) were extracted from the SkaPa register and re-coded to either caries-free (no sign of demineralisation, demineralisation limited to the outer enamel, or fissure sealant) or caries-affected. Surfaces coded with a demineralisation reaching the enamel-dentin junction or extending into the dentin (i.e., equating to ICDAS levels 3 and above including radiographs when the approximal surfaces could not be inspected), secondary caries, restoration, and a tooth with a crown placement or extracted due to caries or for endodontic reasons were coded as caries-affected. Tooth surfaces with ambiguous codes or which were missing for reasons other than caries (extraction for trauma, orthodontic or unknown reasons) were coded as caries-free. Age, sex, living in a socio-economic deprived area as defined by prevalent parental unemployment and high criminality, and dentist-assigned caries risk score (DARS) were extracted from the register. This study uses the terms dmfs (decayed, missing and filled surfaces in the primary dentition) and DMFS (decayed, missing and filled surfaces in the permanent dentition) for summary caries experience scores.

Children who were aged 3, 4, or 5 years at study entry and with at least 1 tooth surface coded as caries-affected were assigned to one of 5 ECC subtypes (Class 1 to 5) using latent class analysis (LCA). The analysis was performed using the software package poLCA in R (version 1.4.1). Children who were caries-free at baseline were treated as a reference group for comparison of subsequent caries experience.

For longitudinal analyses, a minimum of three follow-up visits were required. This included a follow-up visit 2-3 years post baseline for examination of primary teeth, an examination at the age of 6 and an examination reviewing permanent teeth at least 5 years after the 6-year examination.

Subsequent caries experience in the primary dentition was measured as the difference in dmfs scores between the baseline and subsequent visit. A dichotomous outcome was also created reflecting whether children had any new caries (dmfs greater than at baseline) or no new caries (dmfs unchanged since baseline) in the primary dentition.

Caries experience in the permanent dentition was measured using the DMFS score. A dichotomous outcome was created for any caries in the permanent dentition (DMFS ≥1) or no caries in the permanent dentition. All permanent tooth surfaces were considered as caries-free at baseline, given that these teeth were unerupted when the participants entered the study. Any caries lesions in permanent teeth were therefore considered to be manifestations of incident (i.e., “new”) disease.

For some participants, information on the caries risk score as determined by the dentist was available. Where available, this dentist assigned risk score (DARS) was extracted from the first baseline visit, i.e., the same visit that latent class allocations were developed from, and a sensitivity analysis was performed to compare the performance of the DARS and baseline latent class in predicting new caries lesion development. The DARS was scored as no/low, medium, or high risk.

### 2.3 Statistical analyses

Logistic regression was performed to estimate the association between baseline ECC subtype and odds of developing new caries lesions in primary teeth, and odds of developing any caries lesions in permanent teeth. Negative binomial regression was used to estimate the association between baseline ECC subtype and the counts of subsequent caries experience. Models were fitted separately in children aged 3, 4 and 5 years at study entry. Models were adjusted for sex and follow-up age. The results are expressed as odds ratios, representing the odds of any incident disease in the test group divided by the odds of any incident disease in the control group (OR), and incidence rate ratio (IRR), representing expected count of caries-affected surfaces in the test group divided by the expected count of caries-affected surfaces in the control group^15^

Sensitivity analysis was performed using the DARS caries risk assessment tool using the subset of participants with a DARS score available. Comparative statistics were generated including a Vuong analysis to assess the model fit between the ECC subtype and DARS models of prediction. The Vuong analysis is a test of closeness of fit and can be informative about which model is a better description of the observed data.^16^

### 2.4 Ethical approval

The study was approved by the Swedish Ethical Review Authority (Dnr 2019–05031) as well as the directors of each County Council in Sweden. The study followed the Helsinki declaration including that the person with parental responsibility consented to participation at the child’ s regular dental examination.

## 3 RESULTS

In total, 208,112 children aged 3-5 years old at baseline were potentially eligible for inclusion in the study. Latent class modelling was performed on 31,919 children (15.3%) who had evidence of caries at baseline. Latent class modelling was performed on all children who had caries at baseline (including those with no follow-up data available) as this helps improve model fit, therefore reducing the uncertainty in class allocations and reducing regression dilution bias in the subsequent longitudinal analysis.

Of the eligible children, 128,355 had follow-up data for at least 3 further visits spanning at least 5 years of follow-up and were included in the final sample. These children were classified into 5 types of ECC, with differing caries severity between the 5 classes.

A child at baseline who was recorded as caries-free in the latent class calculation is referred to as Class 0. Class 1 refers to caries mainly affecting pits and fissures of molar teeth, Class 2 refers to caries affecting smooth surface anterior teeth, Class 3 has similar distribution of affected surfaces to class 1 but with greater severity, Class 4 has similar distribution of surfaces to class 2 but with greater severity and Class 5 is rampant caries affecting most tooth surfaces. The distribution of caries lesions within the mouth in these different classes is summarized as the proportion of individuals with caries at each tooth surface in each class and presented graphically (Figure 2).

**FIGURE 2.**
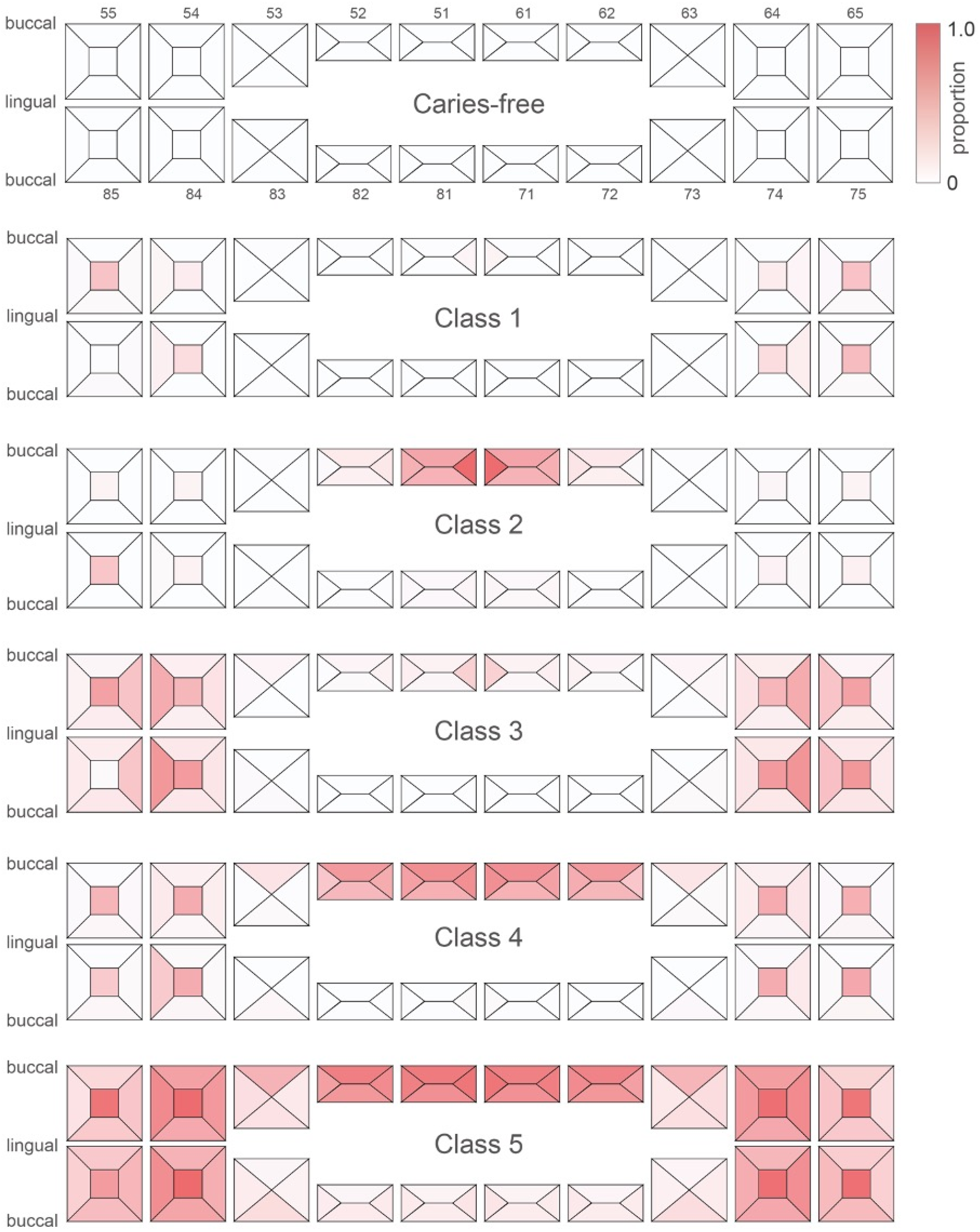
A graphical representation of the 5 ECC classes included in analysis. The scale represents the proportion of individuals in the specific class with caries experience on that surface

At baseline children in Class 1 had a mean dmfs (sd) of 2.7 (1.8), while children in latent Class 5 had a dmfs of 35.5 (14.1) (Table 1).

**TABLE 1.** Baseline demographics of children included in the study

|  | Caries-free | Class 1 | Class 2 | Class 3 | Class 4 | Class 5 | Total |
| --- | --- | --- | --- | --- | --- | --- | --- |
| N (%) children | 103,725 (80.8) | 12,921 (10.1) | 4,595 (3.6) | 3,873 (3.0) | 2,251(1.8) | 990 (0.8) | 128,355 |
| Mean age (sd) years | 4.14 (0.74) | 4.43 (0.64) | 4.29 (0.68) | 4.63 (0.55) | 4.27 (0.66) | 4.43 (0.63) | 4.19 (0.73) |
| Sex (% female) | 49.8 | 49.0 | 44.5 | 43.5 | 46.2 | 40.4 | 49.2 |
| Mean (sd) dmfs | 0 (0) | 2.67 (1.8) | 4.39 (2.5) | 11.30 (5.1) | 14.20 (6.0) | 35.50 (14.1) | 1.29 (4.5) |
| DARS<sup>1</sup> (n) |  |  |  |  |  |  |  |
| no/low risk (score 0) (% of non-missing) | 22,894 (92.5) | 689 (24.9) | 407 (41.7) | 66 (9.0) | 38 (9.2) | 15 (11.2) | 24,109 (80.9) |
| medium risk (score 1) (% of non-missing) | 1,620 (6.5) | 1,167 (42.2) | 309 (31.6) | 179 (24.3) | 110 (26.6) | 30 (22.4) | 3,415 (11.5) |
| high risk (score 2) (% of non-missing) | 248 (1.0) | 907 (32.8) | 261 (26.7) | 491 (66.7) | 266 (64.3) | 89 (66.4) | 2,262 (7.6) |
| missing scores | 78,963 | 10,158 | 3,618 | 3,137 | 1,837 | 856 | 98,569 |
| Deprived area<sup>2</sup> (n) |  |  |  |  |  |  |  |
| 0 (% of non-missing) | 89,478 (95.8) | 11,203 (88.3) | 3,866 (86.9) | 3,172 (83.0) | 1,758 (79.8) | 771 (79.4) | 110,248 (93.8) |
| 1 (% of non-missing) | 449 (0.5) | 172 (1.4) | 67 (1.5) | 89 (2.3) | 57 (2.6) | 24 (2.5) | 858 (0.7) |
| 2 (% of non-missing) | 1588 (1.7) | 494 (3.9) | 188 (4.2) | 200 (5.2) | 142 (6.4) | 72 (7.4) | 2,684 (2.3) |
| 3 (% of non-missing) | 1844 (2.0) | 820 (6.5) | 330 (7.4) | 361 (9.4) | 245 (11.1) | 104 (10.7) | 3,704 (3.2) |
| Missing | 10,366 | 232 | 144 | 51 | 49 | 971 | 10,861 |

1) DARS= Dentist assigned caries risk score using a 3-level scale, no/low risk, medium risk and high risk.
2) A deprived area is a geographically defined district with low socio-economic status and high criminal impact on the local community and is scored as: 0=low deprivation area, 1=deprived area, 2=risk area, 3=highly deprived area.

**Table 2.** Summary of Vuong results for model fit of the latent class against the caries risk model

|  |  | 3-year-old cohort |  |  | 4-year-old cohort |  |  | 5-year-old cohort |  |  |
| --- | --- | --- | --- | --- | --- | --- | --- | --- | --- | --- |
|  |  | Z<br>statistic | p<br>value | Preferred<br>model | Z<br>statistic | p<br>value | Preferred<br>model | Z<br>statistic | p<br>value | Preferred<br>model |
| Odds ratio at<br>follow-up 1 | Raw | 0.44 | 0.33 | Latent Class | 0.13 | 0.45 | Latent Class | 3.11 | <0.01 | Latent Class |
|  | AIC<sup>1</sup> | 0.30 | 0.38 | Latent Class | 0.04 | 0.45 | Latent Class | 3.02 | <0.01 | Latent Class |
|  | BIC<sup>2</sup> | -0.17 | 0.43 | Caries Risk | -0.30 | 0.38 | Caries Risk | 2.67 | <0.01 | Latent Class |
| Incident rate<br>ratio at follow-up 1 | Raw | -2.18 | 0.01 | Caries Risk | 2.77 | <0.01 | Latent Class | -0.51 | 0.31 | Caries Risk |
|  | AIC<sup>1</sup> | -2.43 | <0.01 | Caries Risk | 2.70 | <0.01 | Latent Class | -0.68 | 0.25 | Caries Risk |
|  | BIC<sup>2</sup> | -3.27 | <0.01 | Caries Risk | 2.45 | <0.01 | Latent Class | -1.30 | 0.10 | Caries Risk |
| Odds ratio at<br>follow-up 2 | Raw | 1.42 | 0.08 | Latent Class | 2.85 | <0.01 | Latent Class | 0.58 | 0.28 | Latent Class |
|  | AIC<sup>1</sup> | 0.93 | 0.18 | Latent Class | 2.54 | <0.01 | Latent Class | 0.25 | 0.40 | Latent Class |
|  | BIC<sup>2</sup> | -0.74 | 0.23 | Caries Risk | 1.37 | 0.08 | Latent Class | -0.97 | 0.17 | Caries Risk |
| Incident rate<br>ratio at follow-up 2 | Raw | 1.25 | 0.11 | Latent Class | 3.11 | <0.01 | Latent Class | 0.85 | 0.20 | Latent Class |
|  | AIC<sup>1</sup> | 0.57 | 0.29 | Latent Class | 2.78 | <0.01 | Latent Class | 0.36 | 0.36 | Latent Class |
|  | BIC<sup>2</sup> | -1.76 | 0.04 | Caries Risk | 1.53 | 0.06 | Latent Class | -1.43 | 0.07 | Caries Risk |
| Odds ratio at<br>follow-up 3 | Raw | 0.62 | 0.27 | Latent Class | 1.87 | 0.03 | Latent Class | 3.30 | <0.01 | Latent Class |
|  | AIC<sup>1</sup> | 0.26 | 0.40 | Latent Class | 1.67 | 0.05 | Latent Class | 3.07 | <0.01 | Latent Class |
|  | BIC<sup>2</sup> | -0.98 | 0.16 | Caries Risk | 0.92 | 0.18 | Latent Class | 2.24 | 0.01 | Latent Class |
| Incident rate<br>ratio at follow-up 3 | Raw | 0.47 | 0.32 | Latent Class | 1.87 | 0.03 | Latent Class | 4.07 | <0.01 | Latent Class |
|  | AIC<sup>1</sup> | 0.13 | 0.45 | Latent Class | 1.67 | 0.05 | Latent Class | 3.90 | <0.01 | Latent Class |
|  | BIC<sup>2</sup> | -1.02 | 0.15 | Latent Class | 0.92 | 0.18 | Latent Class | 3.27 | <0.01 | Latent Class |

1. AIC (Akaike information criterion).
2. BIC (Bayesian information criterion).

The mean (sd) follow-up period was 2.9 (1.1) years for primary teeth, and 7.2 (0.6) years for permanent teeth.

In longitudinal analysis, baseline latent class was associated with subsequent incident caries experience during follow-up. Children with ECC at baseline had greater odds and higher incidence rate than children who at baseline were caries free. Among children who had ECC, there were differences in subsequent caries experience in children with different classes of ECC at baseline. Many of the children who had severe ECC at baseline went on to develop caries in the permanent dentition. As an example, 83% of the children who were aged 5 years and had class 5 disease at baseline went on to develop caries in the permanent teeth by age 11, versus 51% of the children who were caries-free at age 5 (adjusted OR 4.90). When looking at the burden of incident disease (i.e., count of tooth surfaces which became caries-affected during follow-up) there were similar associations, where children with ECC had greater incidence rate ratios for caries surfaces than children who were caries-free at baseline. The results are shown graphically in figure 3 and are provided in full within the supplementary material in Table S2.

**FIGURE 3.**
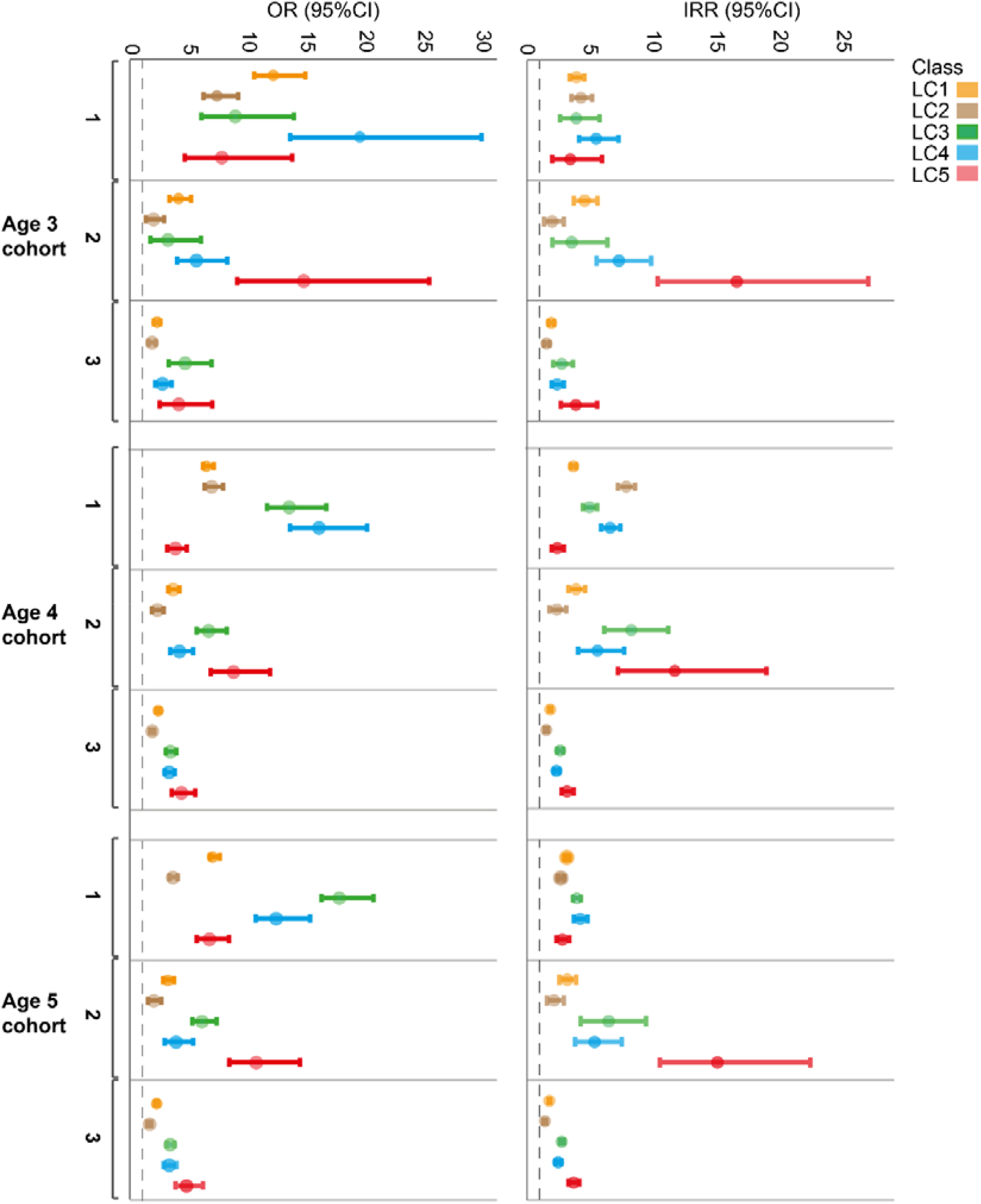
Estimates of association between baseline ECC class and subsequent caries outcomes. Outcome 1=2-3 years post baseline, 2=at age 6, 3=at minimum 5 years post baseline. The plot is split by the association between subtype and odds of incident disease (OR) and between subtype and count of new disease (IRR).

In addition to predicting future disease onset (odds) and severity (count), it is possible that baseline class predicts the spatial distribution of disease within the mouth. To explore this, the distribution of caries in the permanent dentition is summarized by baseline class, in the 5-year-old cohort who were between the ages of 11-13 at their third visit (n=48,433). This group was chosen as an example as there was the largest mean (sd) interval between baseline and visit 3 in years at 7.5 (0.6) years. When examining the distribution of caries-affected surfaces at the final visit, there was a trend for people in the more severe ECC groups to have more caries lesions, and weak evidence for a difference in distribution in caries lesions (Figure 4).

**FIGURE 4.**
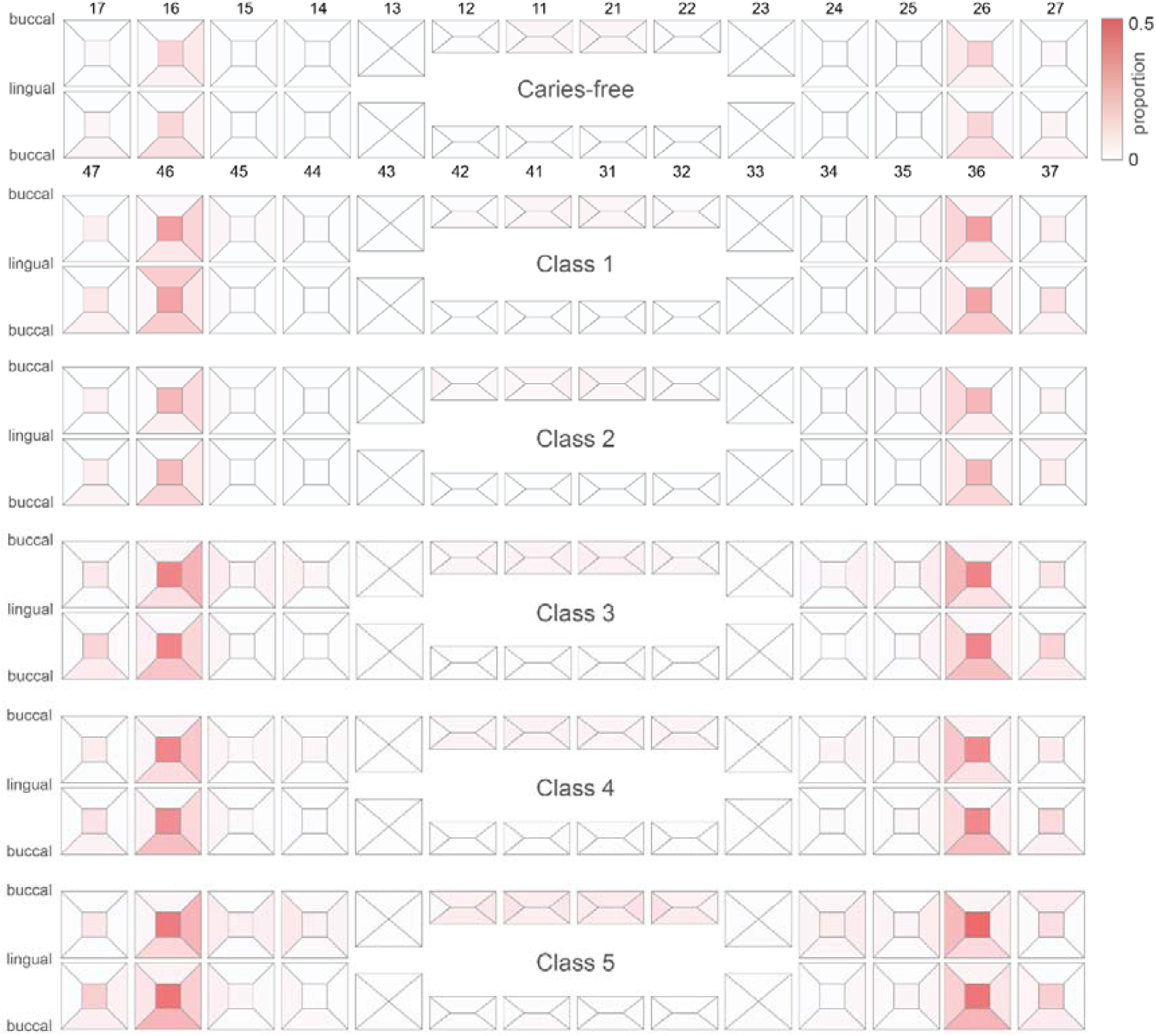
A graphical representation of the 5 ECC classes included in analysis for the children aged 5 years at baseline at their third visit (ages 11 – 13). The scale represents the proportion of individuals in the specific class with caries experience on that surface

For comparison, sensitivity analysis was performed in the subset of participants with a non-missing baseline DARS. This subset included 29,786 children. Logistic and negative binomial regression analyses were performed in this same subset using DARS and latent class as predictors, and model fit compared. In general, the models had similar performance for the age 3 cohort, but latent class provided better prediction than DARS for children aged 4 or 5 years at baseline.

## 5 DISCUSSION

This study tested the hypothesis that children with different patterns of ECC at initial presentation would have differences in subsequent disease experience. The study found that children with all subtypes of ECC at baseline had greater risk for developing caries in permanent teeth than children who were caries-free at baseline, in keeping with existing literature^8^, but demonstrated that this risk differs between groups with different initial presentation. The greatest risk was seen in the ECC subtypes with most severe initial caries presentation. These relationships were stable when comparing children aged 3, 4 and 5 years at entry into the study. This may be because children who have severe ECC at presentation have oral health behaviours,^17^ microbial profiles,^18^ or genetic predisposition,^19^ which place them at high risk for progressive or new disease. Given that the study was nested in a primary care register, the results cannot be explained as a result of inability to access care^20^, but there may be differences in the patterns of health service use or actual care delivered between groups. The results are in keeping with a recent study comparing the risk factors of ECC subtypes, which found distinct biological and environmental risk factors.^9^ Compared to baseline dentist-assigned risk score, ECC subtype performed well in estimating caries risk. This suggests that determining children’s ECC subtypes could be complementary to existing risk assessments that are known to not perform well.^21^

Analysis in the primary dentition is difficult to interpret and this may be due to bias from measurement phenomena. Children with all subtypes of ECC were at high risk of developing further disease in primary teeth, however, there was no clear dose-response relationship between ECC subtype and further caries in primary teeth, and there was variation between different age groups, where children who were older at baseline developed fewer new carious surfaces in comparison to younger children. This may relate to saturation of the dmfs index. As there are a finite number of surfaces that can become carious, children who entered the study with a higher number of affected surfaces (e.g., children in class 5 had almost half of their dentition already affected by caries at baseline) had less potential to develop new disease, creating a bias towards the null which is stronger in children with high caries experience at baseline, i.e., older children and those in the more severe ECC classes. It is also possible that saturation occurs when high-risk surfaces (for example approximal surfaces) develop caries more readily than low-risk surfaces.

In this study 19.2% of children included in the final sample had ECC, versus 15.3% in baseline group. This may indicate a sampling bias, where the final study group is enriched for children at higher risk as these children attend a dentist more regularly for screening or treatment so are more likely to have enough follow-up visits to meet the inclusion criteria. We note that the prevalence of ECC in this study is slightly higher than previously reported (11.4%)^22^. Despite this, the prevalence of ECC in the study remains low in a global context, reflecting that Sweden has one of the lowest prevalences of ECC globally.^23^ Caution is needed when considering other populations where the risk factors and patterns of disease presentation may differ. Regardless of local differences in profile however, it seems likely that a similar approach may yield improvement in caries assessment, but this is not tested in the present study.

The main strengths of the study include the large sample size, nationally representative sampling frame and use of data from clinical examinations. Data from SKaPa have recently been validated in 6- and 12-year-olds, demonstrating satisfactory reliability and validity for the assessment of dental caries.^14^ The main limitation is that data on biological and environmental causes of caries were not available, so we are unable to comment on the underlying mechanisms driving high caries rates in some classes, and suggest this as a topic for future research. For the purposes of prediction however we note that it is not necessary to understand the underlying mechanism.

In summary, the study shows that assignment of ECC subtype at baseline visit can help caries risk prediction and risk assessment, and highlights a potentially new avenue for precision dentistry.^8^ The results from this study further bolster the existing knowledge that past caries experience is a strong predictor for future experience. Finally, we note that ECC remains a prevalent disease even in this cohort in a country with low caries experience and access to comprehensive dental care. This suggests that ECC, while often referred to as preventable, remains an exceptionally difficult disease to prevent at the population level.

## Data Availability

Information on access procedures for the SKaPa register is provided at http://www.skapareg.se/forskning/ (in Swedish).

## ACKNOWLEDGEMENT

The authors thank The Swedish Quality Registry for Caries and Periodontal Diseases (SKaPa) for providing access to the registry data.

## AUTHOR CONTRIBUTION

SH, IJ and KD conceptualized the study, AG, SH and IJ performed formal analyses, PLH and IJ contributed financial support, AG, SH, AE and IJ drafted the manuscript and all authors reviewed, edited, and approved the manuscript prior to final submission.

## SUPPORTING INFORMATION

Additional supporting information may be found in the online version of the article at the publisher’s website.

