## Supporting information for "Subtypes of early childhood caries predict future caries experience"

**Table S1**. Summary of caries outcomes in each class for primary teeth in the (**A**) 3-year-old, (**B**) 4-year-old and (**C**) 5-year-old cohorts

| **(A) 3-year-old cohort** | **Caries-free** | **Class 1** | **Class 2** | **Class 3** | **Class 4** | **Class 5** | **Total** |
| --- | --- | --- | --- | --- | --- | --- | --- |
| Mean age (sd) years at follow-up 1 | 6.45 (0.70) | 6.36 (0.73) | 6.47 (0.68) | 6.51 (0.70) | 6.59 (0.68) | 6.76 (0.54) | 6.45 (0.70) |
| Mean time (sd) years between baseline – follow-up 1 | 3.62 (0.79) | 3.53 (0.82) | 3.65 (0.80) | 3.90 (0.86) | 3.88 (0.79) | 4.08 (0.74) | 3.62 (0.80) |
| Mean dmfs (sd) at follow-up 1 | 1.70 (3.75) | 9.16 (7.15) | 10.20 (8.77) | 12.80 (10.3) | 19.00 (12.0) | 28.90 (19.7) | 2.58 (5.50) |
| Mean dmfs (sd) increment baseline – follow-up 1 | 1.70 (3.75) | 6.89 (6.73) | 7.39 (7.94) | 6.75 (8.20) | 9.25 (8.64) | 6.05 (7.58) | 2.20 (4.52) |
| N (%) with dmfs increment > 0 between baseline and follow-up 1 | 6,773 (30.9) | 898 (85.0) | 466 (77.2) | 114 (80.3) | 243 (89.7) | 57 (77.0) | 8,551 (35.6) |
| Mean age (sd) years at follow-up 2 | 6.00 (0.00) | 6.00 (0.00) | 6.00 (0.00) | 6.00 (0.00) | 6.00 (0.00) | 6.00 (0.00) | 6.00 (0.00) |
| Mean time (sd) years between baseline – follow-up 2 | 3.17 (0.38) | 3.18 (0.38) | 3.19 (0.39) | 3.39 (0.49) | 3.28 (0.45) | 3.32 (0.47) | 3.17 (0.38) |
| Mean dmfs (sd) at follow-up 2 | 0.04 (0.32) | 0.20 (0.70) | 0.09 (0.41) | 0.15 (0.55) | 0.32 (0.96) | 0.68 (1.29) | 0.06 (0.37) |
| N (%) with DMFS >0 at follow-up 2 | 571 (2.61) | 106 (10.0) | 30 (4.97) | 11 (7.75) | 36 (13.3) | 21 (28.4) | 775 (3.22) |
| Mean age (sd) years at follow-up 3 | 9.70 (0.50) | 9.72 (0.49) | 9.70 (0.48) | 9.51 (0.53) | 9.62 (0.53) | 9.64 (0.49) | 9.70 (0.50) |
| Mean time (sd) years between baseline – follow-up 3 | 6.87 (0.36) | 6.90 (0.33) | 6.89 (0.33) | 6.89 (0.35) | 6.91 (0.31) | 6.96 (0.20) | 6.88 (0.36) |
| Mean dmfs (sd) at follow-up 3 | 0.91 (1.67) | 1.85 (2.41) | 1.50 (2.02) | 2.50 (3.17) | 2.22 (2.84) | 3.57 (6.28) | 0.99 (1.80) |
| N (%) with DMFS >0 at follow-up 3 | 8,377 (38.3) | 620 (58.7) | 321 (53.2) | 104 (73.2) | 169 (62.4) | 53 (71.6) | 9,644 (40.1) |

| **(B) 4-year-old cohort** | **Caries-free** | **Class 1** | **Class 2** | **Class 3** | **Class 4** | **Class 5** | **Total** |
| --- | --- | --- | --- | --- | --- | --- | --- |
| Mean age (sd) at follow-up 1 | 6.62 (0.64) | 6.94 (0.30) | 6.65 (0.62) | 6.92 (0.33) | 6.91 (0.36) | 6.96 (0.21) | 6.66 (0.61) |
| Mean time (sd) between baseline – follow-up 1 | 3.18 (0.93) | 3.36 (0.69) | 3.09 (0.89) | 3.37 (0.74) | 3.40 (0.74) | 3.46 (0.68) | 3.20 (0.90) |
| Mean dmfs (sd) at follow-up 1 | 1.33(3.09) | 7.36 (5.81) | 7.42 (6.85) | 15.0 (8.11) | 18.1 (9.66) | 27.2 (17.30) | 2.94 (5.91) |
| Mean dmfs (sd) increment baseline – follow-up 1 | 1.33 (3.09) | 4.79 (5.22) | 4.87 (5.73) | 6.11 (5.78) | 7.27 (7.01) | 3.44 (5.34) | 2.03 (3.98) |
| N (%) with dmfs increment > 0 between baseline and follow-up 1 | 12,689 (27.7) | 4,099 (77.3) | 1,378 (66.4) | 1,003 (86.2) | 972 (87.6) | 283 (68.2) | 20424 (36.6) |
| Mean age (sd) years at follow-up 2 | 6.00 (0.00) | 6.00 (0.00) | 6.00 (0.00) | 6.00 (0.00) | 6.00 (0.00) | 6.00 (0.00) | 6.00 (0.00) |
| Mean time (sd) years between baseline – follow-up 2 | 2.56 (0.66) | 2.42 (0.62) | 2.44 (0.64) | 2.45 (0.66) | 2.49 (0.66) | 2.49 (0.69) | 2.54 (0.65) |
| Mean dmfs (sd) at follow-up 2 | 0.03 (0.25) | 0.12 (0.55) | 0.08 (0.43) | 0.25 (0.84) | 0.17 (0.99) | 0.35 (1.13) | 0.05 (0.36) |
| N (%) with DMFS >0 at follow-up 2 | 927 (2.02) | 376 (7.09) | 93 (4.48) | 141 (12.1) | 89 (8.02) | 64 (15.4) | 1,690 (3.02) |
| Mean age (sd) years at follow-up 3 | 10.30 (0.71) | 10.50 (0.68) | 10.50 (0.70) | 10.40 (0.72) | 10.40 (0.71) | 10.40 (0.74) | 10.35 (0.71) |
| Mean time (sd) years between baseline – follow-up 3 | 6.89 (0.34) | 6.90 (0.33) | 6.89 (0.36) | 6.90 (0.36) | 6.90 (0.35) | 6.88 (0.37) | 6.89 (0.71) |
| Mean dmfs (sd) at follow-up 3 | 0.95(1.70) | 1.94 (2.53) | 1.63 (2.31) | 2.70 (3.72) | 2.39 (2.88) | 3.22 (3.87) | 1.15 (1.99) |
| N (%) with DMFS >0 at follow-up 3 | 18,055 (65.0) | 3,272 (61.7) | 1,148 (55.4) | 809 (69.6) | 762 (68.7) | 308 (74.2) | 24,354 (43.6) |

| **(C) 5-year-old cohort** | **Caries-free** | **Class 1** | **Class 2** | **Class 3** | **Class 4** | **Class 5** | **Total** |
| --- | --- | --- | --- | --- | --- | --- | --- |
| Mean age (sd) years at follow-up 1 | 7.04 (0.86) | 7.01 (0.87) | 7.08 (0.86) | 7.00 (0.87) | 7.23 (0.86) | 7.53 (0.78) | 7.04 (0.86) |
| Mean time (sd) years between baseline – follow-up 1 | 2.05 (0.86) | 2.02 (0.88) | 2.09 (0.87) | 2.02 (0.89) | 2.24 (0.87) | 2.55 (0.80) | 2.05 (0.87) |
| Mean dmfs (sd) at follow-up 1 | 1.21 (2.72) | 6.55 (4.73) | 5.44 (5.44) | 13.90 (6.67) | 15.90 (8.37) | 25.70 (16.50) | 3.29 (5.92) |
| Mean dmfs (sd) increment baseline – follow-up 1 | 1.21 (2.72) | 3.98 (4.18) | 3.40 (4.39) | 4.92 (4.44) | 5.26 (5.28) | 3.54 (4.84) | 1.97 (3.49) |
| N (%) with dmfs increment > 0 between baseline and follow-up 1 | 10,075 (28.0) | 4,834 (73.7) | 1,129 (58.9) | 2,253 (87.7) | 724 (83.2) | 364 (72.7) | 19,379 (40.0) |
| Mean age (sd) years at follow-up 2 | 6.00 (0.00) | 6.00 (0.00) | 6.00 (0.00) | 6.00 (0.00) | 6.00 (0.00) | 6.00 (0.00) | 6.00 (0.00) |
| Mean time (sd) years between baseline – follow-up 2 | 1.01 (0.09) | 1.01 (0.11) | 1.01 (0.10) | 1.01 (0.12) | 1.01 (0.11) | 1.02 (0.15) | 1.01 (0.10) |
| Mean dmfs (sd) at follow-up 2 | 0.03 (0.25) | 0.08 (0.46) | 0.06 (0.39) | 0.16 (0.65) | 0.13 (0.73) | 0.34 (1.30) | 0.05 (0.36) |
| N (%) with DMFS >0 at follow-up 2 | 540 (1.50) | 308 (4.69) | 56 (2.92) | 38 (1.48) | 49 (5.63) | 69 (13.8) | 1,240 (2.56) |
| Mean age (sd) years at follow-up 3 | 12.60 (0.59) | 12.60 (0.59) | 12.60 (0.59) | 12.60 (0.59) | 12.60 (0.59) | 12.60 (0.58) | 12.60 (0.59) |
| Mean time (sd) years between baseline – follow-up 3 | 7.58 (0.58) | 7.61 (0.58) | 7.58 (0.58) | 7.62 (0.58) | 7.64 (0.58) | 7.64 (0.57) | 7.59 (0.58) |
| Mean dmfs (sd) at follow-up 3 | 1.40 (2.20) | 2.69 (3.35) | 2.14 (2.93) | 4.07 (4.54) | 3.70 (4.38) | 5.44 (6.51) | 1.83 (2.84) |
| N (%) with DMFS >0 at follow-up 3 | 18,238 (50.6) | 4,567 (69.6) | 1,192 (62.2) | 1,994 (77.7) | 674 (77.5) | 416 (83.0) | 27,801 (48.5) |

**Table S2**. Summary of OR (new caries) and IRR (severity of new disease) from each follow-up for the (**A**) 3-year-old, (**B**) 4-year-old and (**C**) 5-year-old cohorts

| **(A) 3-yearold cohort** | **Caries-free** | **Class 1** | **p value** | **Class 2** | **p value** | **Class 3** | **p value** | **Class 4** | **p value** | **Class 5** | **p value** |
| --- | --- | --- | --- | --- | --- | --- | --- | --- | --- | --- | --- |
| OR (se) at follow-up 1 | Reference | 12.59 (1.09) | <2E-16 | 7.60 (1.10) | <2E-16 | 9.22 (1.24) | <2E-16 | 20.25 (1.22) | <2E-16 | 8.03 (1.32) | 5.76E-14 |
| IRR (se) at follow-up 1 | Reference | 4.04 (1.07) | <2E-16 | 4.39 (1.10) | <2E-16 | 4.02 (1.21) | 5.08E-13 | 5.60 (1.15) | <2E-16 | 3.59 (1.31) | 1.72E-6 |
| OR (se) at follow-up 2 | Reference | 4.19 (1.12) | <2E-16 | 1.99 (1.21) | 0.000347 | 3.25 (1.37) | 0.000199 | 5.78 (1.20) | <2E-16 | 15.28 (1.30) | <2E-16 |
| IRR (se) at follow-up 2 | Reference | 4.54 (1.19) | <2E-16 | 2.01 (1.28) | 0.00514 | 3.54 (1.60) | 0.00734 | 7.22 (1.38) | 8.99E-10 | 16.40 (1.81) | 2.38E-6 |
| OR (se) at follow-up 3 | Reference | 2.29 (1.07) | <2E-16 | 1.85 (1.09) | 1.46E-13 | 4.79 (1.21) | 2.27E-16 | 2.77 (1.14) | 9.52E-16 | 4.22 (1.30) | 2.64E-8 |
| IRR (se) at follow-up 3 | Reference | 2.05 (1.05) | <2E-16 | 1.66 (1.07) | 1.40E-13 | 2.88 (1.14) | 2.62E-15 | 2.51 (1.10) | <2E-16 | 3.98 (1.20) | 1.68E-14 |

| **(B) 4-year-old cohort** | **Caries-free** | **Class 1** | **p value** | **Class 2** | **p value** | **Class 3** | **p value** | **Class 4** | **p value** | **Class 5** | **p value** |
| --- | --- | --- | --- | --- | --- | --- | --- | --- | --- | --- | --- |
| OR (se) at follow-up 1 | Reference | 6.66 (1.04) | <2E-16 | 7.13 (1.06) | <2E-16 | 13.99 (1.10) | <2E-16 | 16.62 (1.11) | <2E-16 | 3.92 (1.11) | <2E-16 |
| IRR (se) at follow-up 1 | Reference | 3.79 (1.03) | <2E-16 | 7.96 (1.04) | <2E-16 | 5.07 (1.06) | <2E-16 | 6.70 (1.06) | <2E-16 | 2.52 (1.10) | <2E-16 |
| OR (se) at follow-up 2 | Reference | 3.71 (1.07) | <2E-16 | 2.32 (1.12) | 4.01E-14 | 6.85 (1.10) | <2E-16 | 4.28 (1.12) | <2E-16 | 9.06 (1.15) | <2E-16 |
| IRR (se) at follow-up 2 | Reference | 4.00 (1.09) | <2E-16 | 2.50 (1.14) | 8.52E-12 | 8.37 (1.16) | <2E-16 | 5.70 (1.17) | <2E-16 | 11.80 (1.28) | <2E-16 |
| OR (se) at follow-up 3 | Reference | 2.40 (1.03) | <2E-16 | 1.87 (1.05) | <2E-16 | 3.47 (1.07) | <2E-16 | 3.35 (1.07) | <2E-16 | 4.45 (1.12) | <2E-16 |
| IRR (se) at follow-up 3 | Reference | 3.79 (1.03) | <2E-16 | 7.96 (1.04) | <2E-16 | 5.07 (1.06) | <2E-16 | 6.70 (1.06) | <2E-16 | 2.52 (1.10) | <2E-16 |

| **(C) 5-year-old cohort** | **Caries-free** | **Class 1** | **p value** | **Class 2** | **p value** | **Class 3** | **p value** | **Class 4** | **p value** | **Class 5** | **p value** |
| --- | --- | --- | --- | --- | --- | --- | --- | --- | --- | --- | --- |
| OR (se) at follow-up 1 | Reference | 7.21 (1.03) | <2E-16 | 3.70 (1.05) | <2E-16 | 18.43 (1.06) | <2E-16 | 12.84 (1.10) | <2E-16 | 6.94 (1.11) | <2E-16 |
| IRR (se) at follow-up 1 | Reference | 3.28 (1.03) | <2E-16 | 2.80 (1.05) | <2E-16 | 4.06 (1.04) | <2E-16 | 4.33 (1.07) | <2E-16 | 2.92 (1.09) | <2E-16 |
| OR (se) at follow-up 2 | Reference | 3.25 (1.08) | <2E-16 | 2.02 (1.15) | 8.48E-7 | 6.27 (1.09) | <2E-16 | 3.99 (1.17) | <2E-16 | 11.09 (1.15) | <2E-16 |
| IRR (se) at follow-up 2 | Reference | 3.19 (1.11) | <2E-16 | 2.18 (1.20) | 1.79E-05 | 6.42 (1.15) | <2E-16 | 5.33 (1.26) | 6.19E-13 | 14.90 (1.33) | <2E-16 |
| OR (se) at follow-up 3 | Reference | 2.25 (1.03) | <2E-16 | 1.63 (1.05) | <2E-16 | 3.46 (1.05) | <2E-16 | 3.37 (1.09) | <2E-16 | 4.90 (1.13) | <2E-16 |
| IRR (se) at follow-up 3 | Reference | 1.90 (1.02) | <2E-16 | 1.54 (1.03) | <2E-16 | 2.88 (1.03) | <2E-16 | 2.59 (1.05) | <2E-16 | 3.81 (1.06) | <2E-16 |

**Table S3**. Summary of caries outcomes in each caries risk group for primary teeth in the (**A**) 3-year-old, (**B**) 4-year-old and (**C**) 5-year-old cohorts

| **(A) 3-year-old cohort** | **Risk 0** | **Risk 1** | **Risk 2** | **Total** |
| --- | --- | --- | --- | --- |
| Mean age (sd) years at follow-up 1 | 6.44 (0.72) | 6.52 (0.69) | 6.41 (0.71) | 6.44 (0.72) |
| Mean time (sd) years between baseline – follow-up 1 | 3.60 (0.80) | 3.69 (0.80) | 3.58 (0.82) | 3.61 (0.81) |
| Mean dmfs (sd) at follow-up 1 | 1.44 (3.44) | 5.13 (6.99) | 12.20 (10.90) | 2.26 (5.08) |
| Mean dmfs (sd) increment baseline – follow-up 1 | 1.41 (3.37) | 4.34 (6.20) | 8.67 (8.89) | 2.00 (4.43) |
| Mean age (sd) years at follow-up 2 | 6.00 (0.00) | 6.00 (0.00) | 6.00 (0.00) | 6.00 (0.00) |
| Mean time (sd) years between baseline – follow-up 2 | 3.16 (0.37) | 3.18 (0.38) | 3.17 (0.38) | 3.16 (0.37) |
| Mean dmfs (sd) at follow-up 2 | 0.04 (0.30) | 0.09 (0.56) | 0.23 (0.77) | 0.05 (0.37) |
| Mean age (sd) at follow-up 3 | 9.74 (0.48) | 9.72 (0.51) | 9.76 (0.44) | 9.74 (0.48) |
| Mean time (sd) years between baseline – follow-up 3 | 9.74 (0.48) | 9.72 (0.51) | 9.76 (0.44) | 6.90 (0.33) |
| Mean dmfs (sd) at follow-up 3 | 0.82 (1.55) | 1.17 (2.96) | 1.88 (2.48) | 6.89 (0.33) |

| **(B) 4-year-old cohort** | **Risk 0** | **Risk 1** | **Risk 2** | **Total** |
| --- | --- | --- | --- | --- |
| Mean age (sd) years at follow-up 1 | 6.54 (0.67) | 6.81 (0.49) | 6.88 (0.39) | 6.60 (0.64) |
| Mean time (sd) years between baseline – follow-up 1 | 3.03 (0.96) | 3.30 (0.83) | 3.36 (0.78) | 3.09 (0.94) |
| Mean dmfs (sd) at follow-up 1 | 1.27 (3.00) | 5.69 (6.41) | 12.40 (9.65) | 2.68 (3.80) |
| Mean dmfs (sd) increment baseline – follow-up 1 | 1.17 (2.80) | 3.81 (4.84) | 6.43 (6.03) | 1.90 (3.80) |
| Mean age (sd) years at follow-up 2 | 6.00 (0.00) | 6.00 (0.00) | 6.00 (0.00) | 6.00 (0.00) |
| Mean time (sd) years between baseline – follow-up 2 | 2.49 (0.69) | 2.49 (0.66) | 2.48 (0.69) | 2.49 (0.68) |
| Mean dmfs (sd) at follow-up 2 | 0.03 (0.28) | 0.06 (0.40) | 0.16 (0.64) | 0.05 (0.34) |
| Mean age (sd) at follow-up 3 | 10.40 (0.72) | 10.40 (0.69) | 10.40 (0.71) | 10.43 (0.72) |
| Mean time (sd) years between baseline – follow-up 3 | 6.91 (0.30) | 6.93 (0.28) | 6.93 (0.31) | 6.91 (0.30) |
| Mean dmfs (sd) at follow-up 3 | 0.89 (1.66) | 1.50 (2.41) | 2.18 (2.64) | 1.06 (1.89) |

| **(C) 5-year-old cohort** | **Risk 0** | **Risk 1** | **Risk 2** | **Total** |
| --- | --- | --- | --- | --- |
| Mean age (sd) years at follow-up 1 | 7.09 (0.78) | 7.15 (0.84) | 7.05 (0.84) | 7.09 (0.09) |
| Mean time (sd) years between baseline – follow-up 1 | 2.10 (0.78) | 2.16 (0.85) | 2.06 (0.84) | 2.10 (0.80) |
| Mean dmfs (sd) at follow-up 1 | 1.43 (2.96) | 5.94 (6.28) | 12.40 (8.11) | 2.98 (5.35) |
| Mean dmfs (sd) increment baseline – follow-up 1 | 1.27 (2.70) | 3.29 (4.03) | 5.47 (5.07) | 1.90 (3.43) |
| Mean age (sd) years at follow-up 2 | 6.00 (0.00) | 6.00 (0.00) | 6.00 (0.00) | 6.00 (0.00) |
| Mean time (sd) years between baseline – follow-up 2 | 1.01 (0.08) | 1.01 (0.09) | 1.01 (0.10) | 1.00 (0.09) |
| Mean dmfs (sd) at follow-up 2 | 0.03 (0.28) | 0.07 (0.40) | 0.17 (0.72) | 0.05 (0.36) |
| Mean age (sd) at follow-up 3 | 12.50 (0.61) | 12.60 (0.58) | 12.60 (0.56) | 12.53 (0.60) |
| Mean time (sd) years between baseline – follow-up 3 | 7.53 (0.60) | 7.59 (0.58) | 7.62 (0.56) | 7.54 (0.60) |
| Mean dmfs (sd) at follow-up 3 | 1.40 (2.17) | 2.28 (3.12) | 3.48 (4.20) | 7.54 (0.59) |

**Table S4**. Summary of OR (new caries) and IRR (severity of new disease) from each follow-up for (**A**) the 3-year-old, (**B**) the 4-year-old and (**C**) the 5-year-old cohorts by caries risk

| (A) **3-year-old cohort** | **Risk 0** | **Risk 1** | **p value** | **Risk 2** | **p value** |
| --- | --- | --- | --- | --- | --- |
| OR (se) at follow-up 1 | Reference | 3.65 (1.09) | <2E-16 | 11.35 (1.16) | <2E-16 |
| IRR (se) at follow-up 1 | Reference | 3.14 (1.11) | <2E-16 | 6.28 (1.16) | <2E-16 |
| OR (se) at follow-up 2 | Reference | 1.93 (1.24) | 0.00195 | 4.78 (1.23) | 6.33E-14 |
| IRR (se) at follow-up 2 | Reference | 2.31 (1.31) | 0.00212 | 5.79 (1.42) | 5.39E-7 |
| OR (se) at follow-up 3 | Reference | 1.40 (1.09) | 0.000101 | 2.74 (1.13) | <2E-16 |
| IRR (se) at follow-up 3 | Reference | 1.45 (1.08) | 2.21E-6 | 2.32 (1.11) | 1.28E-15 |

| **(B)** **4-year-old cohort** | **Risk 0** | **Risk 1** | **p value** | **Risk 2** | **p value** |
| --- | --- | --- | --- | --- | --- |
| OR (se) at follow-up 1 | Reference | 4.42 (1.07) | <2E-16 | 15.72 (1.11) | <2E-16 |
| IRR (se) at follow-up 1 | Reference | 3.68 (1.06) | <2E-16 | 6.08 (1.07) | <2E-16 |
| OR (se) at follow-up 2 | Reference | 2.00 (1.17) | 1.12E-05 | 4.79 (1.14) | <2E-16 |
| IRR (se) at follow-up 2 | Reference | 2.01 (1.21) | 0.000278 | 5.17 (1.22) | <2E-16 |
| OR (se) at follow-up 3 | Reference | 1.79 (1.06) | <2E-16 | 3.29 (1.07) | <2E-16 |
| IRR (se) at follow-up 3 | Reference | 1.69 (1.05) | <2E-16 | 2.49 (1.06) | <2E-16 |

| **(C)** **5-year-old cohort** | **Risk 0** | **Risk 1** | **p value** | **Risk 2** | **p value** |
| --- | --- | --- | --- | --- | --- |
| OR (se) at follow-up 1 | Reference | 4.12 (1.06) | <2E-16 | 15.61 (1.10) | <2E-16 |
| IRR (se) at follow-up 1 | Reference | 2.62 (1.06) | <2E-16 | 4.33 (1.07) | <2E-16 |
| OR (se) at follow-up 2 | Reference | 2.78 (1.17) | 3.62E-11 | 5.75 (1.15) | <2E-16 |
| IRR (se) at follow-up 2 | Reference | 2.53 (1.22) | 1.93E-6 | 5.95 (1.24) | <2E-16 |
| OR (se) at follow-up 3 | Reference | 1.76 (1.06) | <2E-16 | 2.90 (1.08) | <2E-16 |
| IRR (se) at follow-up 3 | Reference | 1.58 (1.04) | <2E-16 | 2.41 (1.05) | <2E-16 |
